# Chinese Herbal Medicine Treatment for Alzheimer’s Disease Inpatients in China: Alternative or Complement to Conventional Medicine, a Cross-sectional Study

**DOI:** 10.1101/2023.09.19.23295804

**Authors:** Xiaoping Chen, Hao Yan, Stephen Nicholas, Elizabeth Maitland, Zhengwei Huang, Yong Ma, Xuefeng Shi

## Abstract

**Objectives:** Chinese herbal medicine (CHM), a typical type of complementary and alternative medicine (CAM), has been used to treat Alzheimer’s disease (AD) with costs covered by China’s urban basic medical insurance. Previous studies have demonstrated the treatment effectiveness of CHM for intractable disease and CHM’s ability to reduce medical costs. There has been no research exploring the impact of CHM on AD inpatient hospital costs or whether CHM is a complement or alternative to conventional medicine treatments. We compared the medical costs of AD inpatients, CHM users, and non-CHM users to analyze whether CHM has increased or decreased the AD inpatient costs, to assess whether CHM was an alternative or complementary treatment.

**Methods:** Our cross-sectional research was based on a 5% random sample from the 2010 to 2016 Urban Employee Basic Medical Insurance (UEBMI) and Urban Resident Basic Medical Insurance (URBMI) claim data, yielding information on 1507 urban AD inpatients. Wilcoxon rank sum test and chi square test were applied to the medical cost data with an abnormal distribution. To control for confounding factors, such as demographic (age and sex) and medical costs, the influence between CHM costs and conventional medicine costs were analyzed by quantile regression.

**Results:** CHM users accounted for 79.83% (1203/1507) of the total inpatients. The median inpatient cost of CHM users was RMB13293.95 (USD2084.49), which was higher than non-CHM users’ (RMB8428.10/USD1321.53, P<0.001). The median CHM cost was RMB721.00 (USD113.05). Positive correlations between CHM users and CHM costs were found after controlling for confounder variables (Coef.=0.09, P<0.001).

**Conclusion:** During 2010-2016, nearly 80% of AD inpatients in our sample used CHM. Participation in CHM increased the total inpatient costs, pharmacy costs, and conventional medicine costs of AD inpatients over non-CHM users. CHM mainly plays a complementary or less alternative role to conventional medicine for AD treatment.

## 1 Background

There are currently over 55 million people worldwide living with dementia, with 60% of people with dementia living in low- and middle-income countries[1]. In 2022, the global economic costs of dementia are estimated at USD1.3 trillion[2], with USD167.74 billion dementia costs in mainland China, with 50% of those costs accounted for by hospitalization expenses[3]. Alzheimer’s disease (AD) is the most common type of dementia, with over 10 million suffering in mainland China. AD is a neurodegenerative disease with cognitive impairment at its core. As AD progresses, and other comorbidities occur, patients experience memory loss, behavioral and psychological symptoms of dementia (including irritability, hallucinations, delusions, or behavior disorders), gradually loss of basic living capacity, which may progress over 10 years from a mild memory loss condition to complete disability[4,5,6]. In mainland China, Most AD sufferers are concentrated in the central and eastern regions; the incidence increases exponentially as age increases; and AD as a cause of death rose from the tenth cause in 1990 to fifth in 2019 [4].

AD imposes a heavy economic burden on the patient’s families, the hospital system, and the society, which makes an effective treatment desperately needed. Since modern medicine cannot cure AD, experts are increasing turning to complementary and alternative medicine (CAM) as an AD treatment. Following the U.S. National Center for Complementary and Integrative Health (NCCIH), when CAM is used together with conventional medicine, it is labelled ‘complementary’ and when it is used in place of conventional medicine, it is an ‘alternative’[7]. Research on CAM for AD treatment in Japan, Korea, USA, and Canada confirmed the potential anti-inflammatory, memory improving, and neuroprotective effects of CAM[8]. Chinese herbal medicine (CHM), a special type of CAM, has been used for AD treatment for hundreds of years in China[9], and CHM treatments are covered by China’s basic medical insurance[10,11][11].

CHM’s AD treatment is fundamentally different from conventional medicine’s therapeutic assumption of promoting cognition from a molecular biology perspective[12]. The efficacy of CHM treatment has been shown in clinical trials, where molecular biological experiments have demonstrated neuroprotective and antioxidant effects[13]. For example, CHM’s GRAPE formula has been found in a retrospective cohort study to complement conventional medicine therapy to delay cognitive impairment in patients with mild to moderate AD, although the efficacy of CHM usually requires long-term continuous treatment[14]. Studies in Taiwan, China found that more than 40% of dementia patients used traditional Chinese medicine(TCM), with patients with more comorbidities preferring TCM treatment, and with hospitalization costs lower than non-TCM users [15,16]. Ischemic stroke research reported that TCM use increased medical costs, playing a complementary role in treating stroke[17]. It remains inconclusive whether CHM plays a complementary or an alternative role in AD treatment.

Based on the real-world Chinese urban basic medical insurance data, we explore the impact of CHM treatment on AD inpatient costs, especially compared with conventional medicine costs. From a medical treatment cost perspective, we assess whether CHM’s AD treatment was a complementary or an alternative to conventional medical treatment.

## 2 Materials and Methods

### 2.1 Data source

Using a 5% random sample from the 2010 to 2016 Urban Employee Basic Medical Insurance (UEBMI) and Urban Resident Basic Medical Insurance (URBMI) claim data, the China Health Insurance Research Association (CHIRA) collected information on 1507 urban AD inpatients. Data comprised demographic information (sex, age), insurance type, settlement type, hospital type, hospital level, length of hospital stay, frequency of hospital visit, diagnosis code, city type, region, year, total inpatient costs, pharmacy costs, CHM costs and conventional medicine costs.

### 2.2 Parameter interpretation

**Insurance type** Urban basic medical insurance was a social medical insurance established by the Chinese government to ease people’s medical cost burden. Increasing from 430 million members in 2010 to 740 million in 2016[18,19], China’s urban basic medical insurance comprises urban employee basic medical insurance (UEBMI), and urban resident basic medical insurance (URBMI). UEBMI is jointly paid by employees and employers, based on the disposable income of employees. More limited than UEBMI, URBMI covers primary and secondary school students, children, and other nonworking urban residents, with reimbursements related to the resident’s disposable income[20]. And some provinces have also included the rural population in the basic medical insurance, called Urban and Rural Resident Basic Medical Insurance (URRBMI)[21–23].

**Settlement type** Local settlement, trans-city within province settlement, and trans- provincial settlement are different reimbursement types. Medical insurance payments draw on resources at the provincial, county, and city level, with claimants receiving treatment in the city and province in which they are insured called local settlement; for claimants insured in the province, but not the city, are called trans-city province settlement; and claimants insured in another province are called trans-provincial settlement[24].

**Hospital type** Hospitals are classified into general hospitals, containing specialized units, such as internal medicine, surgery, gynecology, pediatrics; specialized hospitals, providing certain diagnosis and treatment services, such as psychiatric specialties, neurological specialties; traditional Chinese medicine hospitals, providing CHM diagnosis and treatment services; and traditional Chinese medicine- integrated hospital, providing both CHM and conventional medicine diagnosis and treatment services[25].

**Hospital level** There are three levels of hospitals in mainland China, comprising primary, secondary, and tertiary levels, with each level providing a wider range and higher quality of personnel and equipment services, medical training, and medical research to local or provincial patients.

**Diagnosis code** G30, based on the International Statistical Classification of Diseases and Related Health Problems, 10th Edition (ICD-10) was the disease code for AD, with subtypes G30.0 (Alzheimer’s disease with early onset, onset usually before the age of 65), G30.1 (Alzheimer’s disease with late onset, onset usually after the age of 65), G30.8 (other Alzheimer’s disease), and G30.9 (Alzheimer’s disease unspecified)[26].

**City type** Based on Chinese 34 provincial-level administrative regions, we classify cities into provincial capital cities, prefecture-level cities, and municipalities directly under the Central Government (Beijing, Tianjin, Shanghai, and Chongqing) [27].

**Region** China divides provincial administrative regions into eastern, central, western, and northeast regions[28]. Eastern region includes Beijing, Tianjin, Hebei, Shanghai, Jiangsu, Zhejiang, Fujian, Shandong, Guangdong and Hainan. Central region includes Shanxi, Anhui, Jiangxi, Henan, Hubei and Hunan. Western region includes Inner mongoria, Guangxi, Choqing, Sichuan, Guizhou, Yunnan, Tibet, Shaanxi, Gansu, Qinghai, Ningxia and Xinjiang. Northeast region includes Liaoning, Jilin and Heilongjiang. Because of the small sample size in the northeast region, it was included in the eastern region in this paper.

**Pharmacy costs** all costs related to pharmacy items, including CHM costs and conventional medicine costs.

**Non-pharmacy costs** Such as examination costs, nursing costs, diagnosis costs, testing costs, bed costs, and medical consumables costs, except for medicine (pharmacy) costs.

**CHM and non-CHM users** When AD inpatients’ incurred any CHM costs, then the inpatient was a CHM user, otherwise, the inpatient was a non-CHM user.

**CHM** Chinese herbal medicine can be divided into herbal medicine and patent medicine. Herbal medicine is extracted (drying, frying) from plants (ginseng, angelica), animals (tortoise shell, gelatin), minerals (gypsum, calamine). Patent medicine comprises the comprehensive integration of different herbal proportions made into decoctions (*Qi Fu Yin)*, tablets (ginkgo leaf pill), or injections (*xueshuantong* injection).

### 2.3 The Sample

The sample comprises 1507 AD hospitalized patients with a discharge time between January 24, 2010 and December 31,2016. It should be stated in advance that although the medical expenses of all samples are recorded, there are still a small part of missing data. For example, one inpatient’s sex, age, and medical expense was recorded in detail, but his settlement type and hospital level were not reported. Therefore, the percentages for many characteristics in Table 1 do not sum to 100%. All costs were based on a the 2013 RMB100=USD15.68 annual average exchange rate.

**Table 1.**
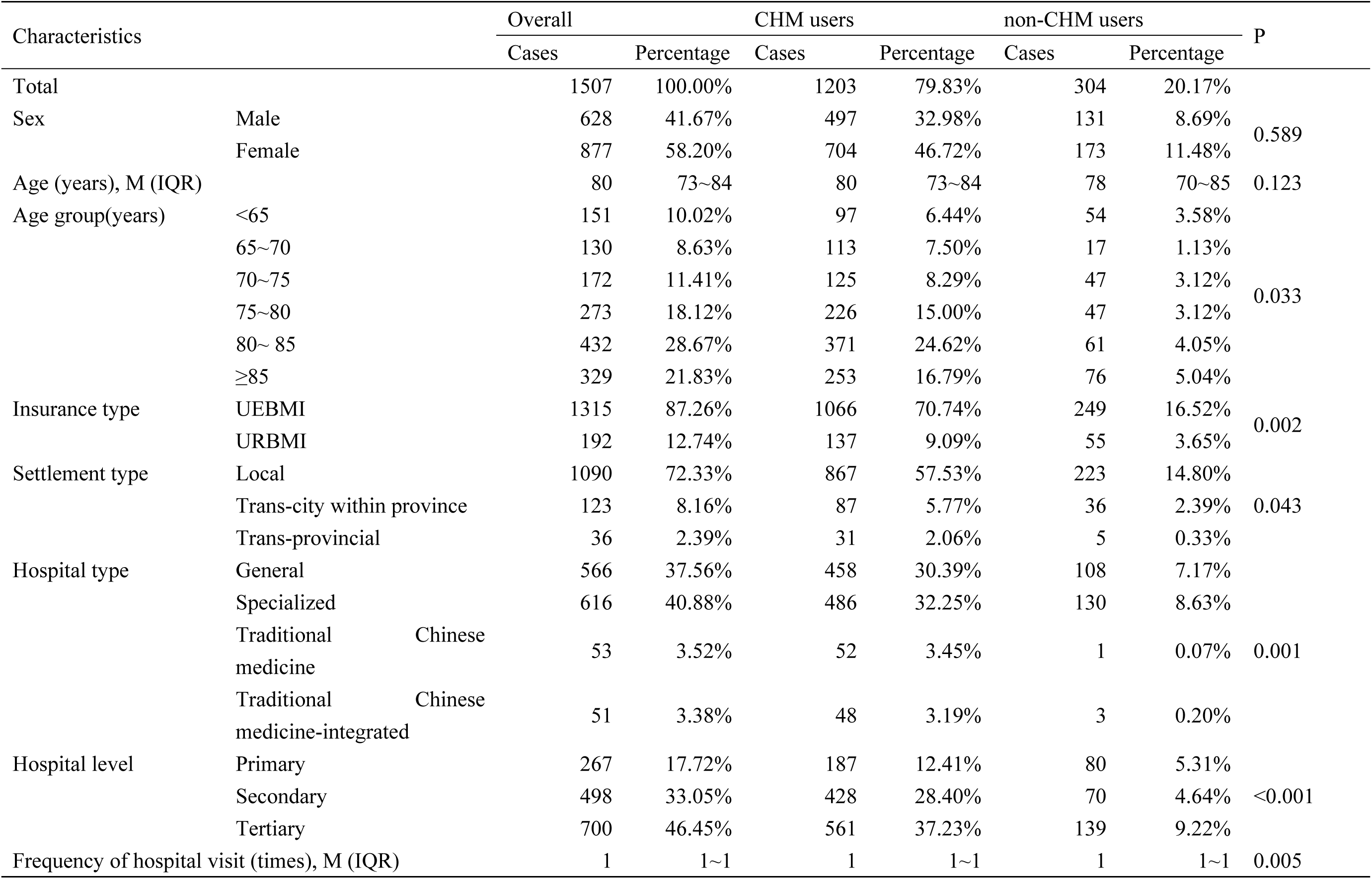
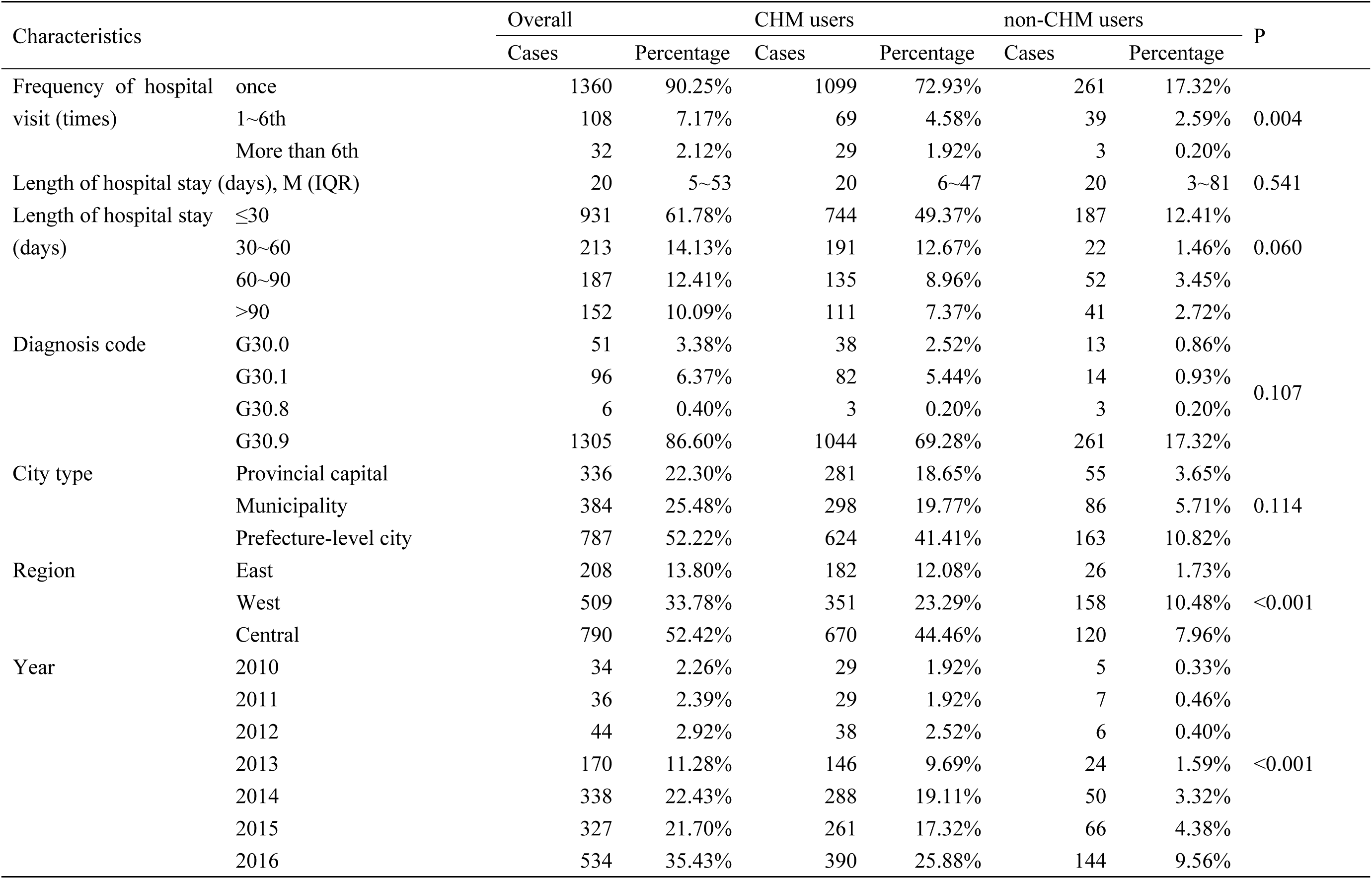
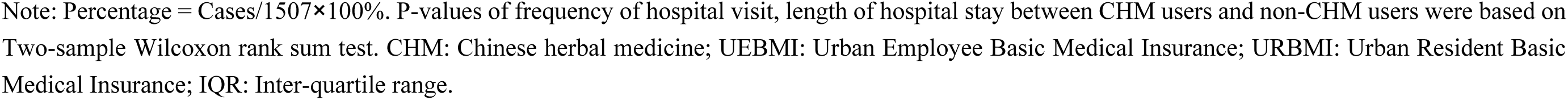
Sample characteristics of CHM users and non-CHM users among AD Inpatients.

| Characteristics |  | Overall |  | CHM users |  | non-CHM users |  | P |
| --- | --- | --- | --- | --- | --- | --- | --- | --- |
|  |  | Cases | Percentage | Cases | Percentage | Cases | Percentage |  |
| Total |  | 1507 | 100.00% | 1203 | 79.83% | 304 | 20.17% |  |
| Sex | Male | 628 | 41.67% | 497 | 32.98% | 131 | 8.69% | 0.589 |
|  | Female | 877 | 58.20% | 704 | 46.72% | 173 | 11.48% |  |
| Age (years), M (IQR) |  | 80 | 73~84 | 80 | 73~84 | 78 | 70~85 | 0.123 |
| Age group(years) | <65 | 151 | 10.02% | 97 | 6.44% | 54 | 3.58% | 0.033 |
|  | 65~70 | 130 | 8.63% | 113 | 7.50% | 17 | 1.13% |  |
|  | 70~75 | 172 | 11.41% | 125 | 8.29% | 47 | 3.12% |  |
|  | 75~80 | 273 | 18.12% | 226 | 15.00% | 47 | 3.12% |  |
|  | 80~ 85 | 432 | 28.67% | 371 | 24.62% | 61 | 4.05% |  |
|  | ≥85 | 329 | 21.83% | 253 | 16.79% | 76 | 5.04% |  |
| Insurance type | UEBMI | 1315 | 87.26% | 1066 | 70.74% | 249 | 16.52% | 0.002 |
|  | URBMI | 192 | 12.74% | 137 | 9.09% | 55 | 3.65% |  |
| Settlement type | Local | 1090 | 72.33% | 867 | 57.53% | 223 | 14.80% | 0.043 |
|  | Trans-city within province | 123 | 8.16% | 87 | 5.77% | 36 | 2.39% |  |
|  | Trans-provincial | 36 | 2.39% | 31 | 2.06% | 5 | 0.33% |  |
| Hospital type | General | 566 | 37.56% | 458 | 30.39% | 108 | 7.17% | 0.001 |
|  | Specialized | 616 | 40.88% | 486 | 32.25% | 130 | 8.63% |  |
|  | Traditional Chinese medicine | 53 | 3.52% | 52 | 3.45% | 1 | 0.07% |  |
|  | Traditional Chinese medicine-integrated | 51 | 3.38% | 48 | 3.19% | 3 | 0.20% |  |
| Hospital level | Primary | 267 | 17.72% | 187 | 12.41% | 80 | 5.31% | <0.001 |
|  | Secondary | 498 | 33.05% | 428 | 28.40% | 70 | 4.64% |  |
|  | Tertiary | 700 | 46.45% | 561 | 37.23% | 139 | 9.22% |  |
| Frequency of hospital visit (times), M (IQR) |  | 1 | 1~1 | 1 | 1~1 | 1 | 1~1 | 0.005 |
|  |  | Cases | Percentage | Cases | Percentage | Cases | Percentage |  |
| Frequency of hospital visit (times) | once | 1360 | 90.25% | 1099 | 72.93% | 261 | 17.32% | 0.004 |
|  | 1~6th | 108 | 7.17% | 69 | 4.58% | 39 | 2.59% |  |
|  | More than 6th | 32 | 2.12% | 29 | 1.92% | 3 | 0.20% |  |
| Length of hospital stay (days), M (IQR) |  | 20 | 5~53 | 20 | 6~47 | 20 | 3~81 | 0.541 |
| Length of hospital stay (days) | ≤30 | 931 | 61.78% | 744 | 49.37% | 187 | 12.41% | 0.060 |
|  | 30~60 | 213 | 14.13% | 191 | 12.67% | 22 | 1.46% |  |
|  | 60~90 | 187 | 12.41% | 135 | 8.96% | 52 | 3.45% |  |
|  | >90 | 152 | 10.09% | 111 | 7.37% | 41 | 2.72% |  |
| Diagnosis code | G30.0 | 51 | 3.38% | 38 | 2.52% | 13 | 0.86% | 0.107 |
|  | G30.1 | 96 | 6.37% | 82 | 5.44% | 14 | 0.93% |  |
|  | G30.8 | 6 | 0.40% | 3 | 0.20% | 3 | 0.20% |  |
|  | G30.9 | 1305 | 86.60% | 1044 | 69.28% | 261 | 17.32% |  |
| City type | Provincial capital | 336 | 22.30% | 281 | 18.65% | 55 | 3.65% | 0.114 |
|  | Municipality | 384 | 25.48% | 298 | 19.77% | 86 | 5.71% |  |
|  | Prefecture-level city | 787 | 52.22% | 624 | 41.41% | 163 | 10.82% |  |
| Region | East | 208 | 13.80% | 182 | 12.08% | 26 | 1.73% | <0.001 |
|  | West | 509 | 33.78% | 351 | 23.29% | 158 | 10.48% |  |
|  | Central | 790 | 52.42% | 670 | 44.46% | 120 | 7.96% |  |
| Year | 2010 | 34 | 2.26% | 29 | 1.92% | 5 | 0.33% | <0.001 |
|  | 2011 | 36 | 2.39% | 29 | 1.92% | 7 | 0.46% |  |
|  | 2012 | 44 | 2.92% | 38 | 2.52% | 6 | 0.40% |  |
|  | 2013 | 170 | 11.28% | 146 | 9.69% | 24 | 1.59% |  |
|  | 2014 | 338 | 22.43% | 288 | 19.11% | 50 | 3.32% |  |
|  | 2015 | 327 | 21.70% | 261 | 17.32% | 66 | 4.38% |  |
|  | 2016 | 534 | 35.43% | 390 | 25.88% | 144 | 9.56% |  |

Note: Percentage = Cases/1507×100%. P-values of frequency of hospital visit, length of hospital stay between CHM users and non-CHM users were based on Two-sample Wilcoxon rank sum test. CHM: Chinese herbal medicine; UEBMI: Urban Employee Basic Medical Insurance; URBMI: Urban Resident Basic Medical Insurance; IQR: Inter-quartile range.

### 2.4 Statistical analysis

Median and inter-quartile range, M (IQR) were used to describe age, length of hospital stay, frequency of hospital visit, total inpatient costs, non-pharmacy costs, pharmacy costs, conventional medicine costs and CHM costs since the Shapiro-Wilk W test revealed they were not normally distributed (P<0.001). Numbers of AD inpatients and proportion of cases were described by cases (n) and proportion (%). The distribution of AD cases between CHM users and non-CHM users were examined by the chi square test and Two-sample Wilcoxon rank sum test, and the differences of age, length of hospital stay, frequency of hospital visit, and costs between CHM users and non-CHM users were analyzed by Two-sample Wilcoxon rank sum test. To quantify the impact of CHM use on total inpatient costs, and the relationship between CHM costs and conventional medicine costs, quantile regression analysis, adjusting for confounding factors, was used. P<0.05 was statistically significant. The statistical processing software was Stata/MP 16.0.

### 2.5 Patient and public involvement

None

## 3 Results

### 3.1 Basics Characteristics

As shown in Table 1, 79.83% (1203/1507) of the 1507 AD inpatients were CHM users, and 20.17% (304/1507) were non-CHM users. The median age was 80 years old, with no statistically significant (P>0.05) between the age of CHM users and non-CHM users, but the age distribution was significant different (P<0.05) between CHM users and non-CHM users. Eighty-seven percent (1315/1507) of AD inpatients were in the UEBMI group, and 70.74% of UEBMI insured were CHM users. There were 72.33% (1090/1507) local settlement inpatients. Only 3.52% (53/1507) of the inpatients were treated at CHM hospital. Most inpatients (90.25%) only visited the hospital once, with CHM users statistically more likely to visit the hospital only once compared to non-CHM users’(P<0.05). On a regional basis, 67.75% (P<0.05) of CHM users came from the west region and central region, which was statistically higher than non-CHM cases. More than half cases (62.31%) of CHM users were treated during the 2014 to 2016 year. Any difference in the distribution of cases among sex, length of hospital stay, diagnosis type, and city type between CHM users and non-CHM users was not statistically significant (P>0.05).

### 3.2 Total Inpatient Costs between CHM Users and non-CHM Users

Table 2 shows the cost difference in CHM users and non-CHM users. The median total inpatient cost for CHM users was RMB13293.95/USD2084.49, significantly higher than non-CHM users (RMB8428.10/USD1321.53, P<0.001). Across sex, age (except 80-85 years old), insurance type, settlement type (local), hospital type (general, specialized), hospital level (secondary, tertiary), frequency of hospital visit (once,1∼6th), length of hospital stay, diagnosis type (G30.9), city type, region, year (2013, 2014, 2015, 2016), CHM users’ total inpatient costs were statistically significant higher than non-CHM users’ (P<0.05).

**Table 2.**
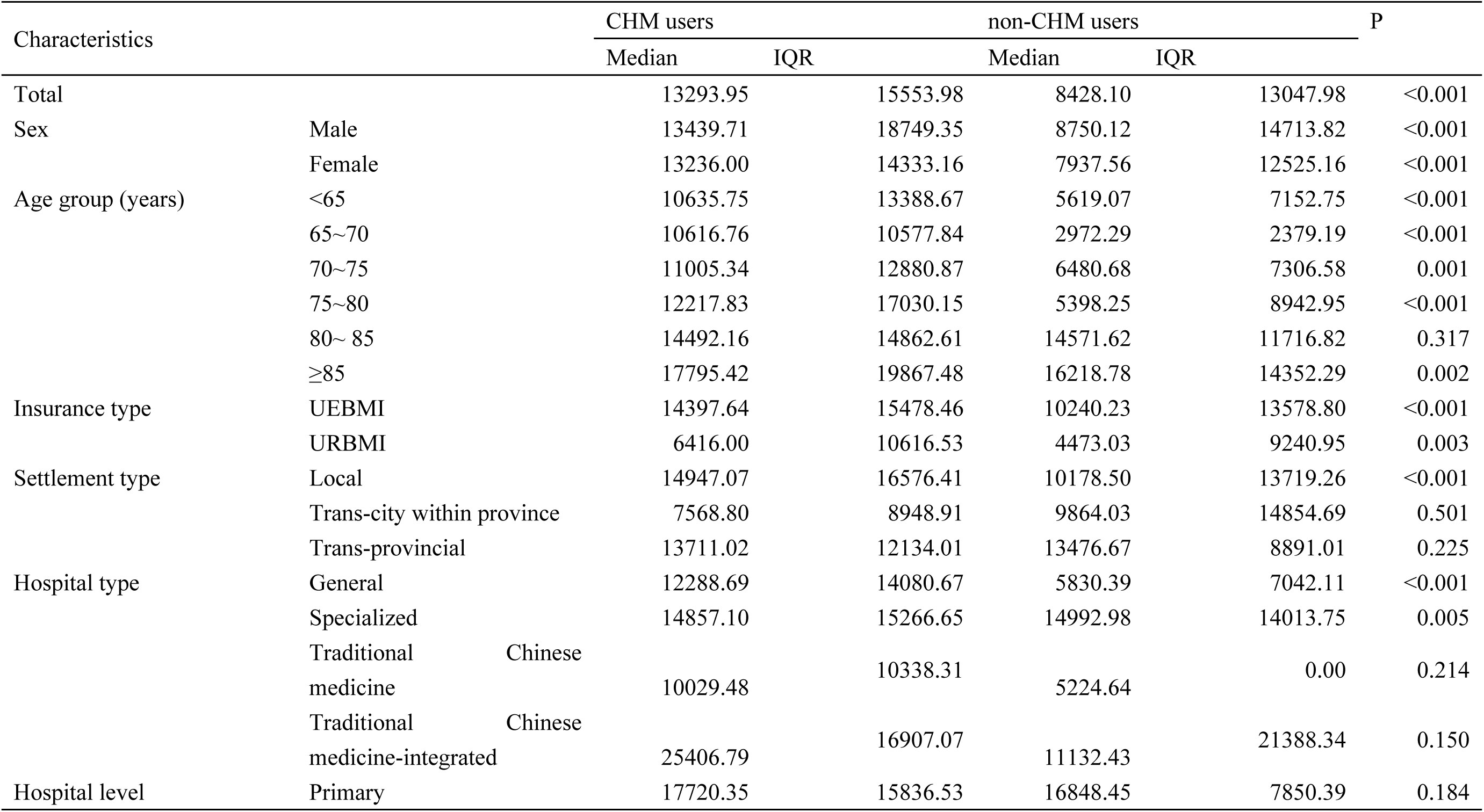
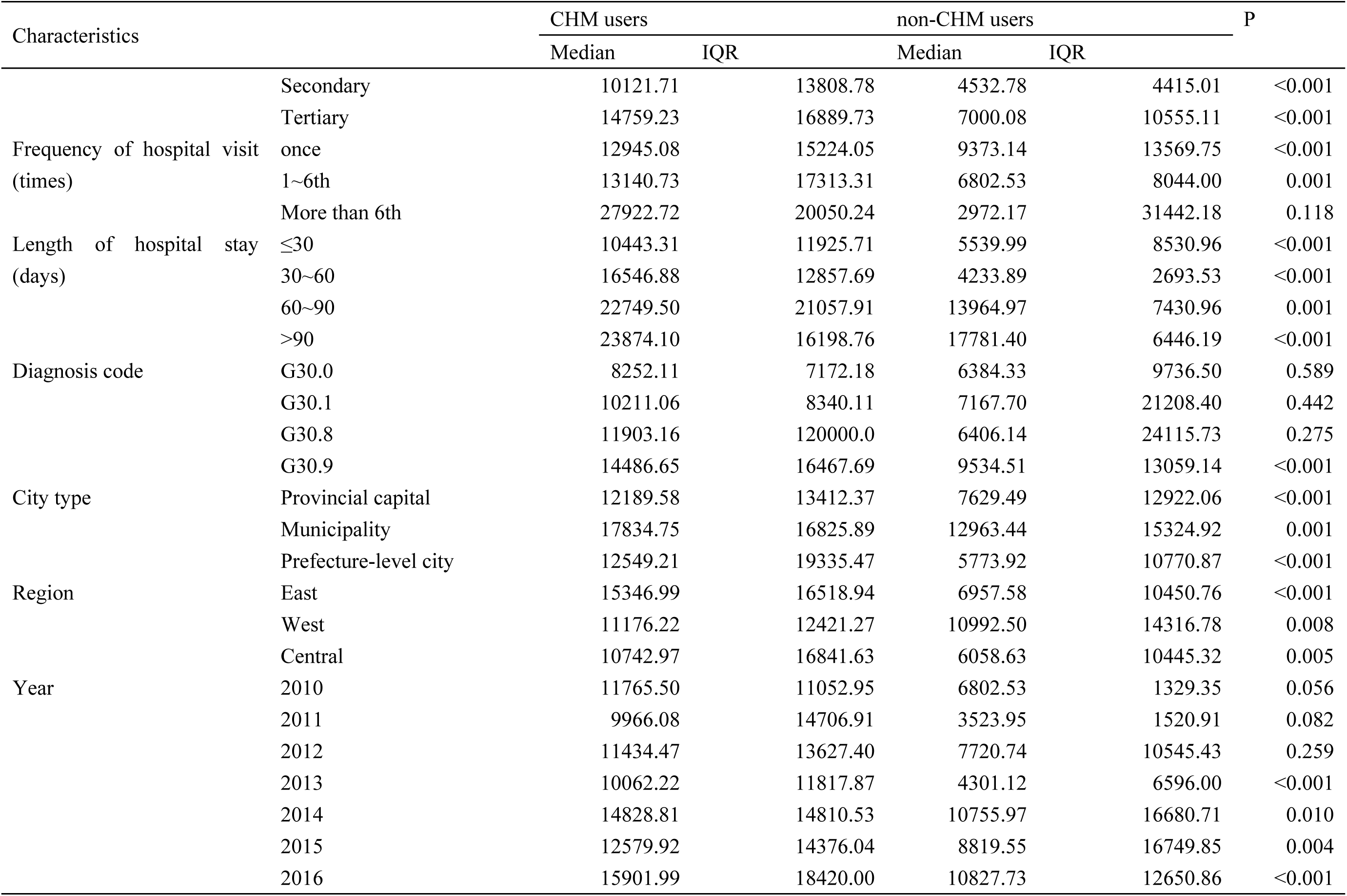
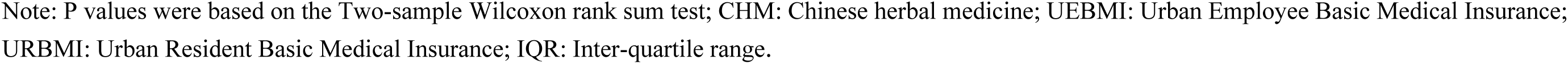
Total Inpatient Costs between CHM Users and non-CHM Users.

### 3.3 Quantile Regression Analysis of Total Inpatient Costs between CHM Users and non-CHM Users

To further analyze the differences in total inpatient costs between CHM users and non-CHM users, we used a quantile regression analysis and included statistically significant factors in Table 2 as confounders. As shown in Table 3, CHM users’ inpatient costs were RMB6898.29/USD1081.65 higher than non-CHM users’ (Coef.=6898.29, P <0.001, R^2^= 0.2133), with confounders (sex, age group, insurance type, settlement type, hospital type, hospital level, frequency of hospital visit, length of hospital stay, type of diagnosis, type of city, region, year) fixed.

**Table 3.** Quantile Regression Analysis of Total Inpatient Costs.

| Characteristics |  | Coef. | Std.Err. | t | P>t | [95% Conf. Interval] |  |
| --- | --- | --- | --- | --- | --- | --- | --- |
| Use of CHM<br>(non-CHM users for reference) | CHM-users | 6898.29 | 1155.914 | 5.97 | 0.000 | 4626.306 | 9170.284 |
| Sex<br>(Male for reference) | Female | 1069.79 | 870.807 | 1.23 | 0.220 | -641.806 | 2781.396 |
| Age group (years)<br>(<65 for reference) | 65~70 | -357.67 | 1514.646 | -0.24 | 0.813 | -3334.760 | 2619.420 |
|  | 70~75 | 1303.02 | 1476.872 | 0.88 | 0.378 | -1599.820 | 4205.870 |
|  | 75~80 | 1890.44 | 1297.271 | 1.46 | 0.146 | -659.392 | 4440.272 |
|  | ≥85 | 2226.26 | 1388.019 | 1.60 | 0.109 | -501.940 | 4954.460 |
| Insurance type<br>(URBMI for reference) | UEBMI | 4393.29 | 1273.408 | 3.45 | 0.001 | 1890.362 | 6896.218 |
| Hospital type<br>(Specialized for reference) | General | -3571.78 | 976.320 | -3.66 | 0.000 | -5490.777 | -1652.793 |
| Hospital level<br>(Secondary for reference) | Tertiary | 2105.62 | 952.754 | 2.21 | 0.028 | 232.949 | 3978.291 |
| Frequency of hospital visit (times)<br>(Once for reference) | 1~6th | 2383.11 | 1536.195 | 1.55 | 0.122 | -636.336 | 5402.556 |
| Length of hospital stay (days)<br>(≤30 for reference) | 30~60 | 4876.86 | 1300.766 | 3.75 | 0.000 | 2320.159 | 7433.561 |
|  | 60~90 | 19573.25 | 1794.292 | 10.91 | 0.000 | 16046.510 | 23100.000 |
|  | >90 | 11781.19 | 1537.534 | 7.66 | 0.000 | 8759.113 | 14803.270 |
| City type<br>(Prefecture-level city for reference) | Provincial capital | -1047.54 | 1140.282 | -0.92 | 0.359 | -3288.805 | 1193.725 |
|  | Municipality | 6311.99 | 1266.385 | 4.98 | 0.000 | 3822.871 | 8801.119 |
| Region<br>(Central for reference) | West | -637.17 | 1189.901 | -0.54 | 0.593 | -2975.963 | 1701.623 |
|  | East | -3147.76 | 1524.595 | -2.06 | 0.040 | -6144.410 | -151.120 |
| Year<br>(2013 for reference) | 2014 | -814.555 | 1823.538 | -0.45 | 0.655 | -4398.783 | 2769.673 |
|  | 2015 | -2833.69 | 1813.871 | -1.56 | 0.119 | -6398.918 | 731.538 |
|  | 2016 | -365.09 | 1770.854 | -0.21 | 0.837 | -3845.766 | 3115.586 |
| _cons |  | 1090.08 | 2548.714 | 0.43 | 0.669 | -3919.507 | 6099.667 |

Note: $R^2 = 0.2133$ in a quantile regression model that was adjusted for use of CHM, sex, age group, insurance type, hospital type, hospital level, frequency of hospital visit, length of hospital stay, city type, region, year. baseline represents the inpatient cost for a 60 to 65 years old male who did not use any CHM with Urban Resident Basic Medical Insurance admitted to a specialized and secondary hospital only once within 30 days' hospital stay in the prefecture-level and central region city in the year of 2013. CHM: Chinese herbal medicine; UEBMI: Urban Employee Basic Medical Insurance; URBMI: Urban Resident Basic Medical Insurance.

### 3.4 Composition of Total Inpatient Costs between CHM Users and non-CHM users

As shown in Table 4, we compared the inpatient cost components between CHM users and non-CHM users, which explained why total inpatient costs were higher in CHM users’. We separated the total inpatient costs into pharmacy costs and non-pharmacy costs for conventional medicine treatment and CHM treatment. Two-sample Wilcoxon rank sum test showed that non-pharmacy costs, pharmacy costs, and conventional medicine costs of CHM users were significantly higher than those of non-CHM users’ (P <0.001).

**Table 4.** Composition of Total Inpatient Costs between CHM Users and non-CHM Users.

| Characteristics | CHM users |  | non-CHM users |  | P |
| --- | --- | --- | --- | --- | --- |
|  | Median | IQR | Median | IQR |  |
| Total inpatient costs | 13293.95 | 15553.98 | 8428.11 | 13047.98 | <0.001 |
| Non-pharmacy costs | 9136.87 | 12093.69 | 5856.95 | 13411.22 | <0.001 |
| Pharmacy costs | 2947.97 | 5416.78 | 672.77 | 2012.79 | <0.001 |
| Conventional medicine costs | 2040.28 | 4263.67 | 672.77 | 2012.79 | <0.001 |
| CHM costs | 721.00 | 1363.92 | -- | -- | -- |

Note: P values were based on the Two-sample Wilcoxon rank sum test. CHM: Chinese herbal medicine; UEBMI: Urban Employee Basic Medical Insurance; URBMI: Urban Resident Basic Medical Insurance; IQR: Inter-quartile range.

### 3.5 Quartile Regression Analysis of CHM Costs

Table 4 shows that CHM treatment had an important impact on pharmacy costs. We conducted a quantile regression on the relationship between conventional medicine costs and CHM costs, adjusted for the impact of confounders (hospital type, hospital level, frequency of hospital visit, length of hospital stay, type of diagnosis, type of city, region, year). As shown in Table 5, conventional medicine costs had a positive impact on CHM costs. When confounders remain unchanged, for every RMB1/USD0.16 increase of conventional medicine costs, CHM costs increased by RMB0.09/USD0.01 (Coef.=0.09, P<0.001, R^2^=0.1998). CHM costs increased with the costs of conventional medicine, indicating that CHM played a complementary role to conventional medicine instead of an alternative treatment role replacing conventional medicine for AD inpatients.

**Table 5.**
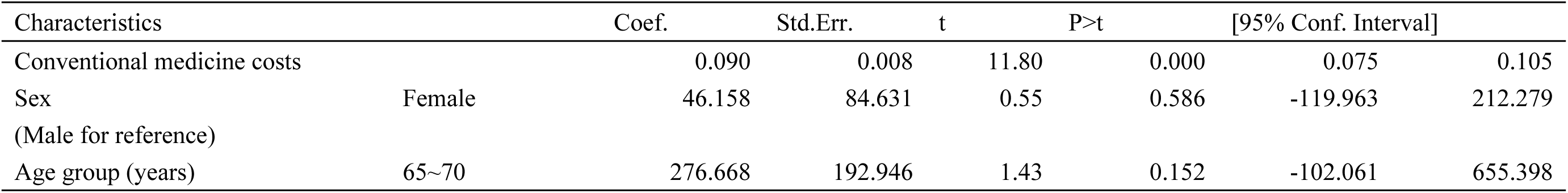
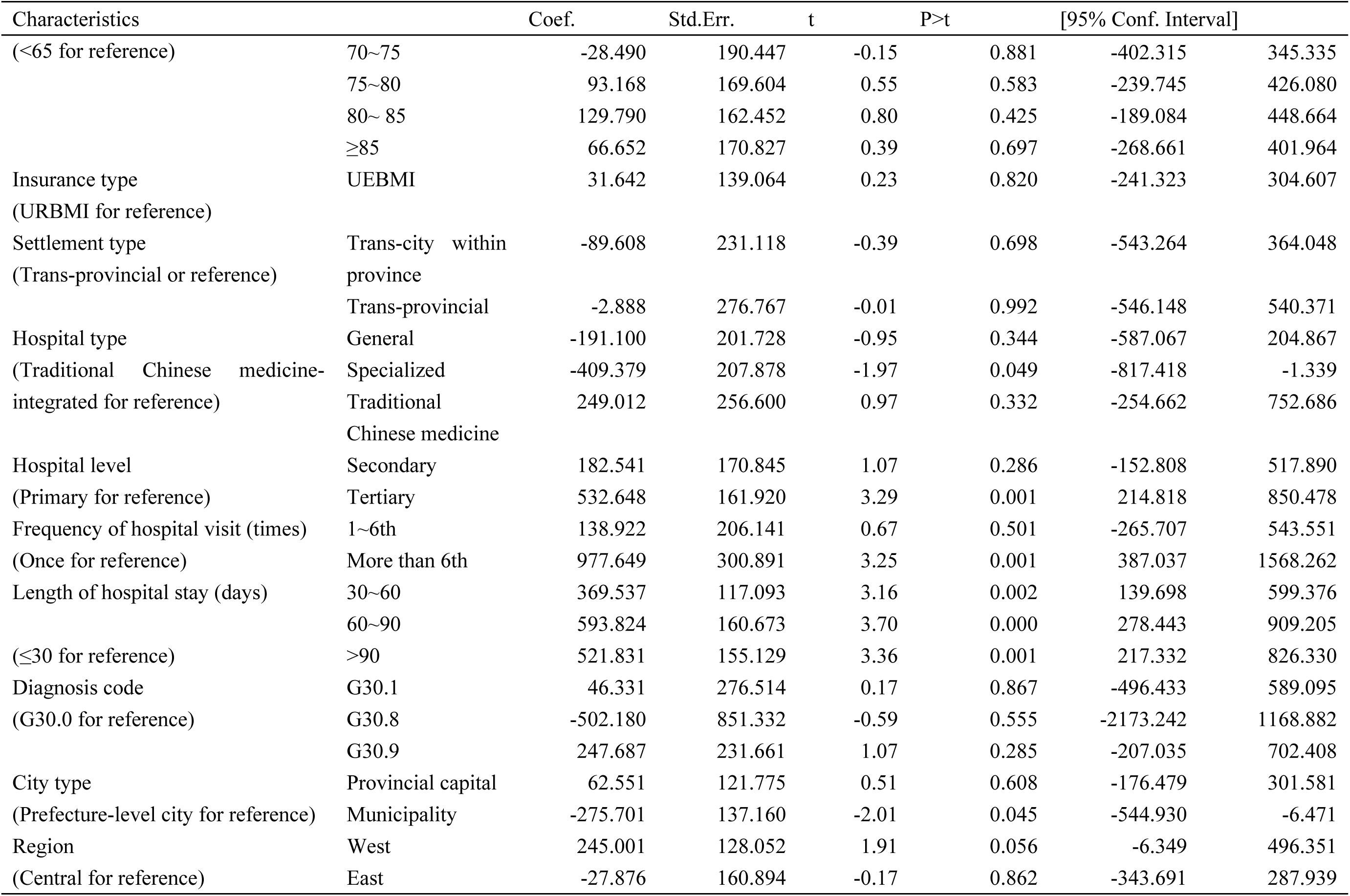
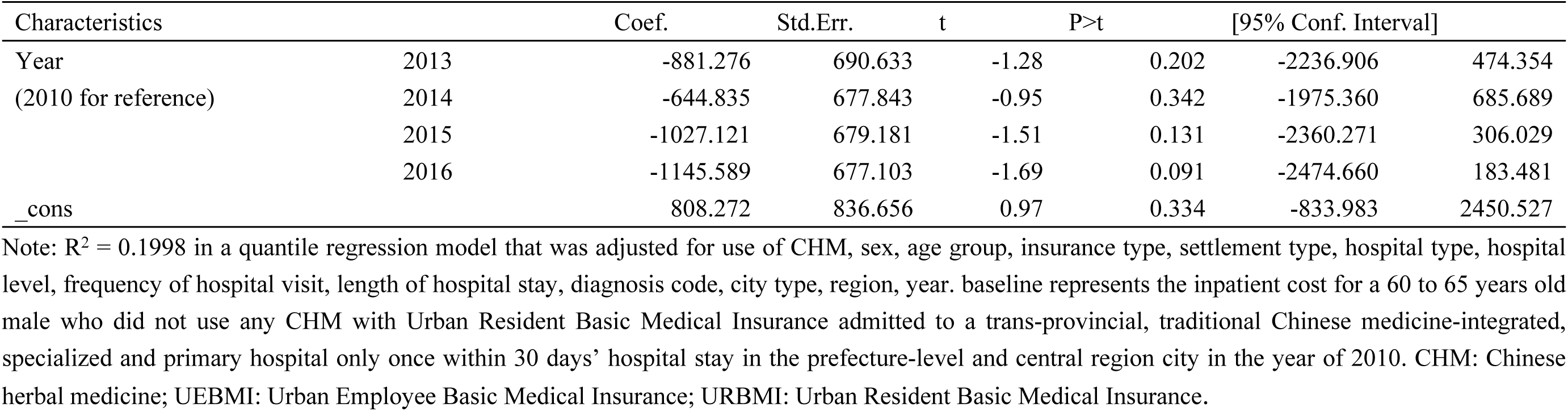
Quartile Regression Analysis of CHM Costs.

## 4 Discussion

CHM is a special type of CAM, and whether CHM is an alternative or complement to conventional medicine remains controversial. Ruling that the effectiveness of herb medicine was based on traditional use rather than scientific research, the UK’s National Health Service medication guidelines do not recommend CHM as supplements to other drugs[29]. In contrast, the U.S. National Center for Complementary and Integrative Health regards herbs as a type of complementary health approach[7]. The Chinese government advocates equal emphasis on CHM and conventional medicine and promotes the mutual complement and coordinated development of both[30]. One approach to explaining whether CHM is a complementary or an alternative treatment process to conventional medicine is from the perspective of medical expenses. Research based on National Health Insurance (NHI) in Taiwan, China has shown that medical costs for uterine fibroid patients with Chinese medicine therapy were lower than Chinese medicine nonusers, and concluded that Chinese medicine was an alternative therapy to conventional medicine[31]. Evidence from mainland China found that the inpatient stroke costs of TCM users were higher than those of TCM nonusers, concluding that TCM was a complementary treatment to conventional medicine[17]. Based on 2010-2016 cross-sectional urban basic medical insurance inpatient cost data in mainland China, we explored whether CHM treatment of AD was an alternative or complement to conventional medical treatment. Compared with non-CHM users (20.17%), 79.83% of AD inpatients received CHM treatment. Two-sample Wilcoxon rank sum test for costs revealed that the non-pharmacy costs, pharmacy costs, and conventional medicine costs of CHM users were statistically significantly higher than those of non-CHM users. As shown in the quantile regression of the relationship between conventional medicine costs and CHM costs, which was adjusted for confounders (including hospital type, hospital level, frequency of hospital visit, length of hospital stay, diagnosis type, city type, region, year), CHM costs increased with rising conventional medicine costs. CHM treatment neither reduced non-pharmacy costs nor pharmacy costs, but acted as additional treatment cost. We conclude that CHM was a complementary therapy rather than an alternative for AD hospitalization treatment.

We suggest that the following reasons help explain the complementary role of CHM in AD treatment. First, conventional medicine dominates the medical market in mainland China. Although China has been exploring CHM treatments for AD for many years, it was not until 2018 that standardized CHM clinical pathways for AD were formally determined[32,33]. In contrast, conventional medicine treatments, such as donepezil and rivastigmine, have been used in medical institutions as anti-AD drugs since 2009[10]. Second, some research has shown that the combination of CHM and conventional medicine can produce better effects than monotherapy. Different from the precise positioning of disease etiology, pathology, and anatomy of conventional medicine, CHM paid attention to the human body and nature environment from a systemic perspective. The combination therapy of CHM and conventional medicine can treat AD from two different perspectives, macroscopic and microscopic, and the effect will definitely be better[12,14]. Third, the profits of drug mark-ups affected doctors’ prescription behavior. Before 2017, several provinces in mainland China implemented a drug mark-up policy, where medicine sold by the hospital was priced higher than the market retail price, providing a profit-making income for the hospital[34]. Finally, the best integrated clinic pathways between conventional medicine and CHM has not been found. To realize the best practice of CHM and conventional medicine, the Chinese government plans to train high-level integrated CHM and conventional medicine experts and general practitioners over the next 10-15 years[35].

## 5 Conclusion

In mainland China, approximately 80% of patients with AD were CHM users. Participation in CHM AD treatment increased the total inpatient costs, pharmacy costs, and conventional medicine costs of AD inpatients over non-CHM users. CHM mainly plays a complementary or less alternative role to conventional medicine for AD treatment.

## Strengths and Limitations

This study has some limitations. Our data were stratified sample from urban basic medical insurance in mainland China with a sampling frequency of one year and one time, which can only sample sectional inpatient costs for AD among its decades disease progression. Furthermore, the severity and comorbidity of AD were important indicators affecting inpatient costs, but these data were not available in our database. Acupuncture and moxibustion were essential non-pharmacy TCM therapies for AD, but data on such treatments were not available. Finally, our data were from 2010 to 2016, but in 2017, the Chinese government proposed a new policy to further promote TCM development, which might influence our conclusions.

## Data Availability

The data that support the findings of this study are available from China Health Insurance Research Association, but restrictions apply to the availability of these data, which were used under license for the current study and so are not publicly available. Data are however available from Xuefeng Shi upon reasonable request and with permission of the China Health Insurance Research Association. If someone wants to request the data from this study, please contact the China Health Insurance Research Association.

## Abbreviations

AD: Alzheimer’s Disease
TCM: Traditional Chinese medicine
CHM: Chinese herbal medicine
UEBMI: Urban Employee Basic Medical Insurance
URBMI: Urban Resident Basic Medical Insurance
URRBMI: Urban and Rural Resident Basic Medical Insurance
IQR: Inter-quartile range

## Declarations

### Ethics approval and consent to participate

Since the claim data the authors used were an anonymized and secondary database, human participants are not directly involved in the study. The Ethics Committee of Dongzhimen Hospital Affiliated to Beijing University of Chinese Medicine (no.2019BZHYLL0201) has waived the subsequent ethical application and informed consent application.

### Consent for publication

Not Applicable.

### Availability of data and materials

The data that support the findings of this study are available from China Health Insurance Research Association, but restrictions apply to the availability of these data, which were used under license for the current study and so are not publicly available. Data are however available from Xuefeng Shi upon reasonable request and with permission of the China Health Insurance Research Association.

If someone wants to request the data from this study, please contact the China Health Insurance Research Association.

### Authors’ Contributions

Xiaoping Chen and Hao Yan developed the research aims, drafted the manuscript, and analyzed the data; Xuefeng Shi, Stephen Nicholas, and Elizabeth Maitland revised the manuscript; Xuefeng Shi and Yong Ma developed the research idea; Yong Ma and Zhengwei Huang oversaw the data collection; Xuefeng Shi collected the data and helped develop the idea and edited the manuscript. All authors reviewed the manuscript.

## Acknowledgments

The authors acknowledge the China Health Insurance Research Association for providing access to their claims data.

